# Elevated urine BMP phospholipids in LRRK2 and VPS35 mutation carriers with and without Parkinson’s disease

**DOI:** 10.1101/2022.08.03.22278207

**Authors:** Sara Gomes, Alicia Garrido, Francesca Tonelli, Donina Obiang, Eduardo Tolosa, Maria Jose Marti, Javier Ruiz-Martinez, Ana Vinagre Aragon, Haizea Hernandez-Eguiazu, Ioana Croitoru, Vicky L Marshall, Theresa Koenig, Christoph Hotzy, Frank Hsieh, Marianna Sakalosh, Elizabeth Tengstrand, Shalini Padmanabhan, Kalpana Merchant, Christof Bruecke, Walter Pirker, Alexander Zimprich, Esther Sammler

## Abstract

Elevated urine bis(monoacylglycerol)phosphate (BMP) levels have been found in gain-of-kinase function LRRK2 G2019S mutation carriers. Here, we have expanded urine BMP analysis to other Parkinson’s disease (PD) associated mutations and found them to be consistently elevated in carriers of LRRK2 G2019S and R1441G/C as well as VPS35 D620N mutations. Urine BMP levels are promising biomarkers for patient stratification and potentially target engagement in clinical trials of emerging targeted PD therapies.

---

Parkinson’s disease (PD) is the most common neurodegenerative movement disorder affecting about 6 million people worldwide with numbers expected to rise and no cure. While the aetiology of most PD cases is poorly understood (e.g. idiopathic), rarer monogenetic forms have advanced our understanding of PD and the development of disease-modifying treatments that are currently being evaluated in clinical trials^1^. One such example is the leucine-rich repeat kinase 2 (LRRK2) and small molecule LRRK2 kinase inhibitors^1^. LRRK2 encodes a large multidomain protein but only 7 PD associated variants have so far been recognized as clearly disease causing^2^. The consensus is that LRRK2 mutations result in gain-of-kinase function, which is mirrored by increased LRRK2 dependent substrate phosphorylation, including Rab10 at threonine 73^2^. However, LRRK2 kinase activation may have much broader application as an underlying disease mechanism; for example in carriers of additional LRRK2 variants of unknown clinical significance^3^, in idiopathic PD^4^ and carriers of the PD causing VPS35 D620N mutation^5^. As such, there is a need for robust and accessible markers for mechanistic disease stratification: urine mass spectrometry studies have revealed a lysosomal dysregulation signature^6^ and elevated bis(monoacylglycero)phosphate (BMP) levels^7^ in carriers of the common LRRK2 G2019S mutation. The latter is consistent with previous observations of decreased BMPs in urine of *lrrk2* knockout mice, and non-human primates treated with LRRK2 kinase inhibitors^8^. It has also been reported that treatment with the LRRK2 inhibitor DNL201 resulted in a marked decrease in urine BMP levels in 122 healthy volunteers and in 28 iPD patients in phase 1 and phase 1b clinical trials, respectively^9^. BMPs belong to a broad group of atypical, negatively charged phospholipids important for membrane formation and function in the endosome-lysosome compartment, including intraluminal vesicles and extracellularly released exosomes^10^.

Here, we have expanded urine BMP analysis to other PD associated mutations that significantly hyperactivate the LRRK2 kinase with greater effect than LRRK2 G2019S: carriers of LRRK2 R1441 hotspot mutations and VPS35 D620N that activates LRRK2 kinase activity by a yet unknown mechanism, and also GBA variant carriers and one individual with a novel homozygous ATP13A2 variant.

22 healthy controls, 31 individuals with iPD, and the following carriers of heterozygous pathogenic variants were included in this study: 18 with LRRK2 G2019S mutation (11 PD/7 non-manifesting carriers (NMC)), 13 with LRRK2 R1441G/C mutation (7 PD/6 NMC) and 10 with the pathogenic VPS35 D620N mutation (9 PD/1 NMC), as well as 10 individuals with PD associated with various GBA risk variants. Additionally, one participant with atypical young onset PD and a novel homozygous ATP13A2 G38D mutation was included. Table 1 gives an overview of participants’ demographics, clinical status with regards to PD and genotype according to groups.

**Table 1.** Demographic information and urine BMP levels in study participants.

|  | Control | iPD | LRRK2<br>G2019S | LRRK2<br>R1441G/C | VPS35<br>D620N | GBA | p-value |
| --- | --- | --- | --- | --- | --- | --- | --- |
|  | N = 22 | N = 31 | N = 18 | N = 13 | N = 10 | N = 10 |  |
| **Age** | 55 | 65.5 | 59.5 | 65 | 62.5 | 61 |  |
| median (range) | (24, 71) | (45, 88) | (42, 81) | (50, 83) | (29, 76) | (49, 64) | <0.001<sup>1</sup> |
| **Sex** |  |  |  |  |  |  |  |
| female/male | 11/11 | 15/16 | 11/7 | 6/7 | 7/3 | 3/7 | 0.5<sup>2</sup> |
| **PD status** |  |  |  |  |  |  |  |
| PD/NMC | -- | -- | 11/7 | 7/6 | 9/1 | 10/0 | -- |
| **Disease duration** |  |  |  |  |  |  |  |
| median (range) | -- | 6<br>(1, 17) | 6<br>(1, 22) | 9<br>(6, 20) | 15<br>(1, 21) | 5<br>(1, 21) | 0.2<sup>1</sup> |
| **Total di-18:1-BMP** | 3.66 | 6.48* | 13.43**** | 9.57** | 15.97**** | 2.56 |  |
| median (range) | (0.72, 8.54) | (1.48, 38.88) | (3.30, 42.56) | (2.70, 34.68) | (6.71, 55.37) | (0.80, 45.53) | <0.001<sup>1</sup> |
| **2,2'-di-18:1-BMP** | 2.04 | 4.58** | 11.59**** | 4.68** | 13.69**** | 1.58 |  |
| median (range) | (0.52, 5.60) | (1.01, 34.58) | (2.86, 33.59) | (1.55, 29.59) | (4.54, 46.10) | (0.59, 21.59) | <0.001<sup>1</sup> |
| **Total 22:6-BMP** | 8.33 | 12.84 | 43.52**** | 28.11*** | 29.19*** | 9.66 |  |
| median (range) | (2.18, 41.14) | (3.28, 104.92) | (6.98, 321.98) | (7.94, 81.34) | (15.51, 162.36) | (3.97, 34.21) | <0.001<sup>1</sup> |
| **2,2'-di-22:6-BMP** | 4.83 | 9.86* | 33.43**** | 21.27*** | 22.61*** | 5.93 |  |
| median (range) | (1.47, 19.59) | (2.17, 60.15) | (5.61, 153.74) | (4.35, 47.35) | (8.98, 126.35) | (2.81, 21.31) | <0.001<sup>1</sup> |

Values are median of the measured BMP normalized to the creatinine amount (ng/mg creatinine).
<sup>1</sup>Kruskal-Wallis rank sum test.
<sup>2</sup>Fisher's exact test.
Values significantly different from Control group: \*p < 0.05; \*\*p < 0.01; \*\*\*p < 0.001, \*\*\*\*p < 0.0001, Dunn's multiple comparison test.
NMC: non-manifesting carrier.
GBA group includes carriers of the following variants: c.762-1G>C, het.del-from exon3, p.Pro138Leufs\*62, p.Thr408Met, p.Ser212\* and p.Thr408Met, p.Glu365Lys, p.Asn409Ser.

Table 1 and Figure 1 show the data for the total and 2,2’-isoforms of the di-18:1-BMP and di-22:6-BMP species that had previously been reported to most strongly discriminate between LRRK2 G2019S mutation carriers and non-carriers^7^. All these BMP isoforms were significantly raised in the LRRK2 G2019S mutation carrier group when compared to controls. A similarly significant increase in BMP levels compared to controls was also seen in the VPS35 D620N and LRRK2 R1441G/C mutation carrier groups (Figure 1). No statistically significant difference compared to controls was observed for the GBA risk variant group. With regards to iPD compared to controls, a statistically significant increase was found for the total and 2,2’-isoforms of di-18:1-BMP and 2,2’-di-22:6-BMP but not total di-22:6-BMPs. We did not observe a statistical difference in these 4 species in GBA variant carriers with PD compared to controls. Data for all other BMP isoforms (Supplementary table 2 and Supplementary figure 1) and all possible group comparisons (Supplementary table 3). Although there is a significant difference in age between the experimental groups (Table 1), the statistically significant differences in BMP levels were maintained when corrected for age differences (Supplementary file 1).

**Figure 1.**
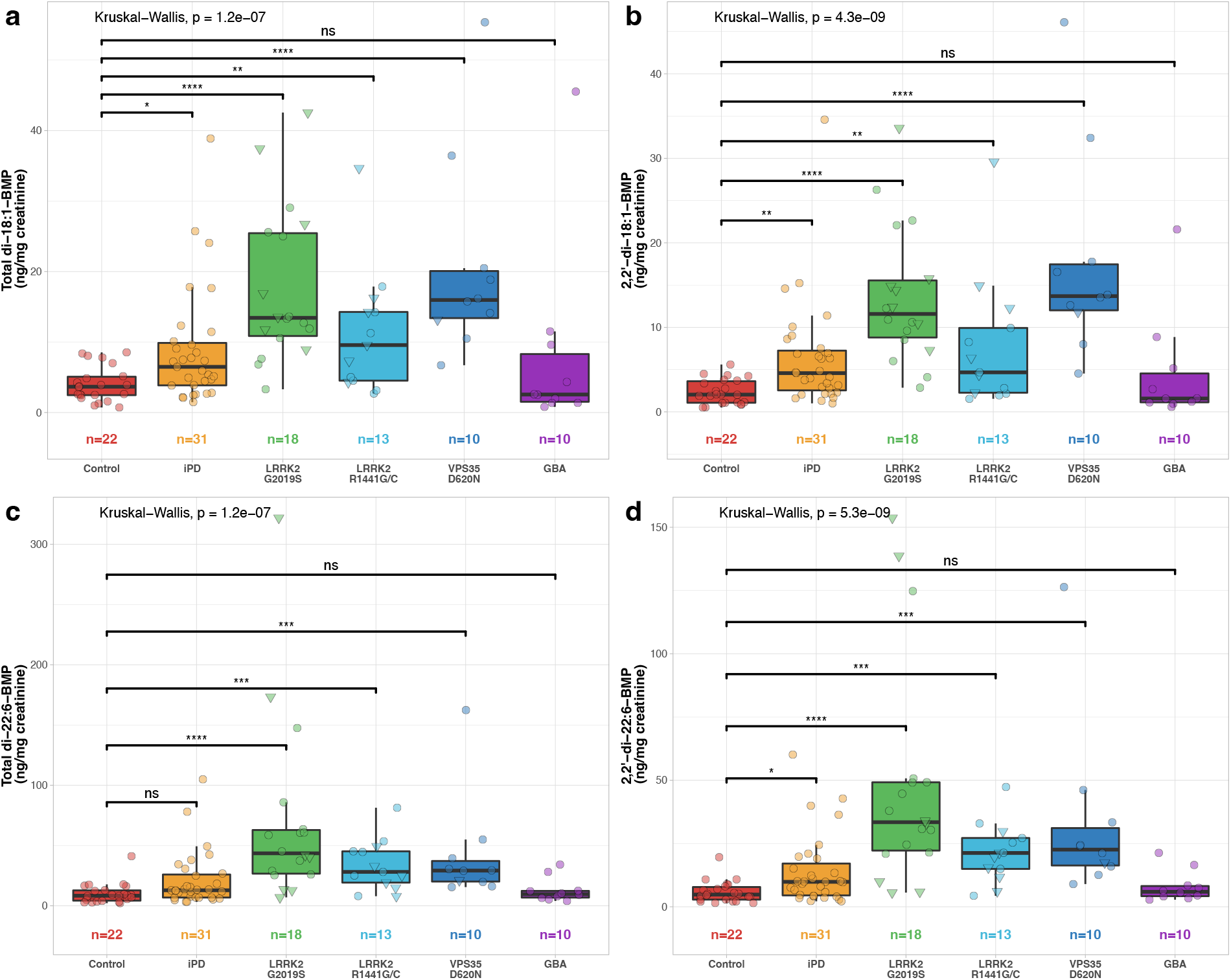
Levels of urine BMP isoforms. Urine levels of **(a)** 2,2’-di-18:1-BMP, **(b)** total di-18:1-BMP, **(c)** 2,2’-di-22:6-BMP, and **(d)** total di-22:6-BMP expressed for all groups. Triangle datapoint shapes indicate non-manifesting mutation carriers. Statistically significant differences between groups were assessed with Kruskal-Wallis test. Post-hoc Dunn’s multiple comparison test was employed to identify groups significantly different from control (\**p* < 0.05, \*\**p* < 0.01, \*\*\**p* < 0.001, \*\*\*\**p* < 0.0001).

BMP levels of the ATP13A2 mutation carrier are not included in the grouped analyses, but all BMP isoforms were high, in particular the total and 2,2’-di-18:1 isoforms, which were more than 3-fold higher than the upper range of the LRRK2 G2019S group (Supplementary table 1).

Using non-parametric using Kruskal Wallis testing, there were no significant differences between PD manifesting and non-manifesting carriers for LRRK2 G2019S and R1441 hotspot mutation carriers for any of the BMP isoforms (Supplementary table 4). Given that there was only one non-manifesting carrier amongst the VPS35 D620N group and all GBA risk variant carriers had a diagnosis of PD, impact of clinical PD status was not assessed for these groups.

At a time of rising cases of PD and emerging disease-modifying treatments, reliable biomarkers that reflect underlying disease mechanisms and have potential for target engagement studies are urgently needed. We and others have highlighted the utility of LRRK2 substrate phosphorylation, e.g., LRRK2-dependent Rab10 phosphorylation at threonine 73, in peripheral blood as a biomarker for LRRK2 kinase activity^5,11,12^. Urine BMPs may be a complementary and in case of LRRK2 G2019S more sensitive biomarker for LRRK2 dysfunction with the additional advantage of being non-invasive and much easier to process than peripheral blood^5,13^.

Our data together with previous results^7,9^ indicates that elevated urine BMP levels are a strong indicator of elevated LRRK2 kinase pathway activity. We observed a significant increase in all BMP isoforms measured for LRRK2 G2019S mutation carriers irrespective of PD status but did not see a significant difference between PD manifesting compared to non-manifesting LRRK2 G2019S carriers which may be due to the relatively small group size (11 PD/7 NMC). For example, the previous study was only able to observe a statistically significant increase in BMP levels in LRRK2 G2019S carriers with PD after combining the results of 2 cohorts (45 PD/36 NMC) but not separately^7^. Additionally, we found a significant elevation in all BMP species measured for carriers of the LRRK2 R1441G/C as well as VPS35 D620N mutations when compared to controls. Importantly, our results provide further evidence that kinase-activating mutations in LRRK2 and the VPS35 D620N mutation operate in a shared pathway regulating endosomal-lysosomal and Golgi-sorting processes that result in elevated urine BMPs in humans^14^. While we did not observe a statistical effect of GBA variant carrier status in comparison to controls, this will need to be assessed further in much larger datasets. The participant with ATP13A2 associated Kufor-Rabek syndrome displayed significantly elevated urine BMP levels. ATP13A2 encodes a transmembrane endolysosomal ATPase that transports polyamines into the cell regulating endolysosomal cargo-sorting and proteostasis through an interaction with the BMP precursor phosphatidylinositol^15^. A finding that contrasted with the previous study^7^ was that urine BMP levels were increased in our iPD group compared to controls and we hypothesize that this is a reflection of the heterogeneity amongst individuals with iPD in itself, which highlights the need for mechanistic stratification of PD patients.

In conclusion, we provide confirmation that urine BMP isoforms are robustly elevated in carriers of the LRRK2 G2019S mutation and provide first time evidence that urine BMPs are also elevated in LRRK2 R1441G/C and VPS35 D620N mutation carriers, consistent with shared biological pathways involved in endolysosomal trafficking. Our results also underlines that there are discriminatory biomarkers for mechanistic patient stratification that may in the future assist in matching patients with the best available targeted treatments.

## Methods

104 participants across 4 different sites were recruited for this study between 2019 and 2021. Controls were self-reported in general good health, did not have a diagnosis or family history of PD or tremor and did not suffer from any other neurodegenerative conditions. All participants gave written informed consent, and the study was approved by the respective local ethics committees. Up to 10 mL of fresh midstream urine was centrifuged for 15 minutes at 2500g and 4°C in most cases within half an hour from collection. The supernatant was transferred, aliquoted into labelled tubes, immediately snap frozen and maintained at −80°C for storage and shipment, until analysis at Nextcea, Inc.

A multiplexed UPLC-MS/MS method was used to simultaneously quantitate molecular species of BMPs in urine, performed by Nextcea, Inc. (Woburn, MA) as before^7^. Normalization, calibration, and data processing was performed as described^7^.

All data analysis was carried out in R (version 4.1.2) and the code is provided in the supplementary information (Supplementary File 1). Non-parametric statistical tests (Kruskal-Wallis with post-hoc Dunn’s test) were used to compare the different groups defined as control, iPD, LRRK2 G2019S, LRRK2 R1441G/C, VPS35 D620N, and GBA. Adjusted *p* values are reported (*p* values lower than 0.001 are reported as *p* < 0.001), and *p* < 0.05 was considered statistically significant. To assess the potential impact of sex and age as predictors of variance of BMP levels, generalised linear models were applied.

## Data Availability

All data produced in the present work are included in the manuscript as supplementary files.

## Data availability

The full BMP and clinical datasets are available in supplementary table 1.

## Acknowledgements

The work was supported by funding from the Michael J. Fox Foundation for Parkinson’s Research [MJFF-020316, MJFF-009262]. ES was supported by a Chief Scientist Office Scottish Senior Clinical Academic Fellowship. We thank all patients and volunteers who have participated in this study. We acknowledge the excellent technical support of the MRC-Protein Phosphorylation and Ubiquitylation Unit (PPU) and MRC PPU Reagents and Services teams. We thank the Data Analysis Group at the School of Life Sciences at the University of Dundee for their useful discussion and guidance.

## CRediT (Contributor Roles Taxonomy) author contributions

S. Gomes: investigation, data curation, formal analysis, writing of original draft, visualization. A. Garrido: investigation, data curation, review and editing of manuscript. F Tonelli: investigation, review and editing of manuscript. D Obiang: investigation. E Tolosa, MJ Marti, J Ruiz-Martinez: resources, review and editing of manuscript. A Vinagre-Aragon, H Hernandez-Eguiazu, I Croitoru: investigation, review and editing of manuscript. V Marshall: resources, review and editing of manuscript. T. Koenig: investigation, review and editing of manuscript. C Hotzy: investigation, review and editing of manuscript. F Hsieh: methodology, investigation, formal analysis, resources, data curation, review and editing of manuscript. M Sakalosh: investigation, formal analysis, review and editing of manuscript. E. Tengstrand: investigation, formal analysis, validation, review and editing of manuscript. S Padmanabhan: conceptualization, review and editing of manuscript. K Merchant: conceptualization, review and editing of manuscript. C Brücke: resources, data curation, review and editing of manuscript, W Pirker: resources, investigation, data curation, review and editing of manuscript. A Zimprich: investigation, resources, data curation, review and editing of manuscript. EM Sammler: conceptualization, data curation, validation, visualization, formal analysis, supervision, project administration, funding acquisition, writing of original draft, review and editing of manuscript.

## Supplementary Material Legends

**Supplementary file 1. R code used for statistical analyses of data**.

**Supplementary figure 1. Levels of urine BMP isoforms for the extended panel**. Urine levels of **(a)** BMP-18:1/22:6, **(b)** BMP-18:2, **(c)** BMP-18:1/18:2, **(d)** BMP-18:2/22:6, **(e)** 2,3’-BMP-18:1, **(f)** 2,3’-BMP-22:6, **(g)** 3,3’-BMP-22:6, **(h)** BMP-20:4/22:6, **(i)** BMP-18:2/20:4, **(j)** BMP-16:0/18:2, **(k)** 3,3’-BMP-18:1, **(l)** BMP-16:0/18:1, **(m)** Lyso-BMP-22:6, **(n)** Lyso-BMP-18:1, **(o)** BMP-20:3/22:6, **(p)** BMP-18:0/20:4, expressed as ng of BMP-per mg of creatinine, are plotted per experimental group (indicated in the x-axis), in boxplots. Individual datapoints are shown, with triangle shapes indicating non-manifesting mutation carriers. Group size is indicated. Statistically significant differences between experimental groups were assessed with Kruskal-Wallis test, and overall p-values are displayed on the top left corner of each plot. Post-hoc Dunn’s multiple comparison test was employed to identify groups significantly different from control (\**p* < 0.05, \*\**p* < 0.01, \*\*\**p* < 0.001, \*\*\*\**p* < 0.0001), except in cases where overall *p* > 0.05 or group size ≤ 2.

**Supplementary table 1. Participant demographic and clinical characteristics**. For each participant, sample collection site is provided as: BCN (Barcelona), VIE (Vienna), DND (Dundee), or SSB (San Sebastian). Also provided are age at study participation, age at PD diagnosis (where applicable), sex (M, for male, and F, for female), experimental group (control, iPD – idiopathic PD –, LRRK2 G2019S, LRRK2 R1441G/C, VPS35 D620N, GBA, or other), and PD status (NMC for non-manifesting mutation carriers, or PD). Other details including additional mutations are listed in the comments column, where applicable. Values for all measured BMP species presented as ng of BMP per mg of creatinine are provided. BQL designates BMP levels that were below quantification level and NM designates values that were not measured for a particular individual.

**Supplementary table 2. Levels of urine BMPs for the extended panel**.

**Supplementary table 3. Multiple group comparison for main urine BMP isoforms**. Summary of the adjusted p-values and significance level for the comparisons between the 5 experimental groups (control, iPD – idiopathic PD –, LRRK2 G2019S, LRRK2 R1441G/C, VPS35 D620N, and GBA), for the four main urine BMP species. Kruskal-Wallis test, with post-hoc Dunn’s multiple comparison test (\**p* < 0.05, \*\**p* < 0.01, \*\*\**p* < 0.001, \*\*\*\**p* < 0.0001, ns: non-significant, *p* > 0.05).

**Supplementary table 4. Comparison between PD-manifesting and non-manifesting LRRK2 mutation carriers for main urine BMP isoforms**. Summary of the adjusted p-values and significance level for the comparison between PD-manifesting (PD) and non-manifesting (NMC) carriers of LRRK2 G2019S or LRRK2 R1441G/C mutations, for the four main urine BMP species. Kruskal-Wallis test, with post-hoc Dunn’s multiple comparison test (ns: non-significant, *p* > 0.05).

