## Supplementary figures and images for "Elevated urine BMP phospholipids in LRRK2 and VPS35 mutation carriers with and without Parkinson’s disease"

### Supplementary Figure 1

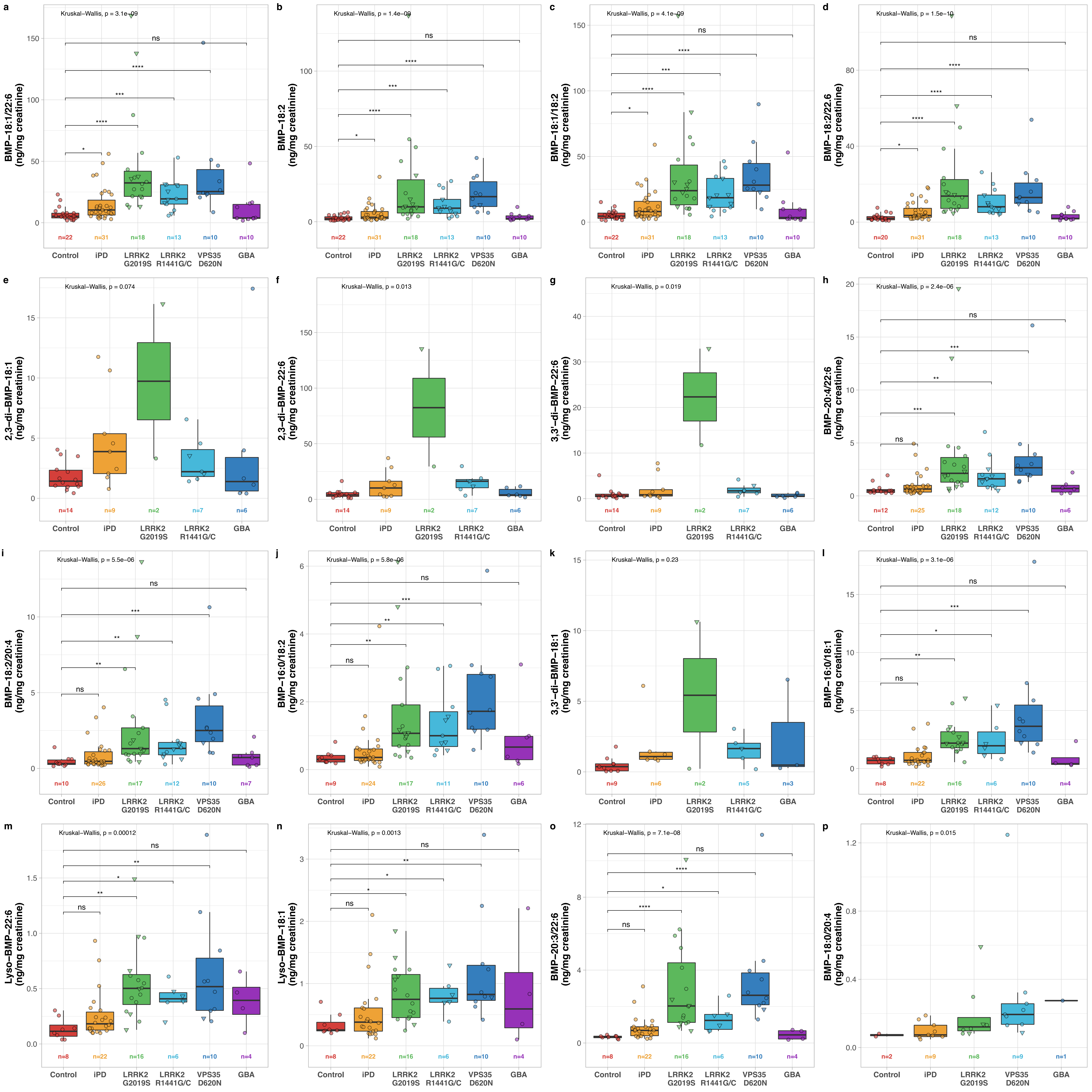
