## Supplementary File 1 for "Elevated urine BMP phospholipids in LRRK2 and VPS35 mutation carriers with and without Parkinson’s disease"

Elevated urine BMP phospholipids in LRRK2 and VPS35 mutation carriers with and without Parkinson's disease

**4 Session Information** **22**

### 1 Dataset

The dataset used for the majority of analyses is the provided **Supplementary Table 1**, saved as `.csv`. To generate **Table 1** the raw dataset was used, which includes all participants' ages at time of study participation. This dataset is not included to comply with journal requirements regarding participant anonymity.

```
# Raw data includes individual participants' ages for analysis. This dataset is not
included in the paper, to comply with journal requirements.
df01 <- read_csv(here::here("raw_data.csv"), col_types =
"cfififffddddddddddddddddddd")
df01 <- clean_names(df01)

# Imported data is provided supplementary table 1.
df02 <- read_csv(here::here("suppl_table_1.csv"), col_types =
"cfffifffddddddddddddddddddd")
df02 <- clean_names(df02)
bmp_variables <- names(df02)[9:28]
bmp_labels <- c("Total di-18:1-BMP",
                "2,2'-di-18:1-BMP",
                "Total di-22:6-BMP",
                "2,2'-di-22:6-BMP",
                "BMP 18:1/22:6",
                "BMP 18:2",
                "BMP 18:1/18:2",
                "BMP 18:2/22:6",
```

```
                 "2,3'-BMP 18:1",
                 "2,3'-BMP 22:6",
                 "3,3'-BMP 22:6",
                 "BMP 20:4/22:6",
                 "BMP 18:2/20:4",
                 "BMP 16:0/18:2",
                 "3,3'-BMP 18:1",
                 "BMP 16:0/18:1",
                 "Lyso-BMP 22:6",
                 "Lyso-BMP 18:1",
                 "BMP 20:3/22:6",
                 "BMP 18:0/20:4")

bmp_vars <- as.list(setNames(bmp_labels, bmp_variables))
```

### 2 Exploratory data analysis

After data checking and cleaning, exploratory analysis was carried out. Data were checked for normal distribution (**Section 2.1**) by plotting density plots (**Figure 1**) and q-q plots (**Figure 2**). Descriptive statistics (**Section 2.2**) were calculated and summarised in **Tables 1** and **2**. Grouped boxplots generated for visualisation are shown in **Figure 3** (**Section 2.3**).

#### 2.1 Distribution

The density plots (**Figure 1**) and q-q plots (**Figure 2**) show that data is not normally distributed, thus non-parametric tests were used for statistical analyses (**Section 3**).

```
dens_list = list()
for (i in names(bmp_vars)) {
  dens_list[[i]] = ggplot(df02, aes(x = !!sym(i))) +
    geom_density(alpha = 0.6,
                 size = 0.25)+
    xlab(bmp_vars[[i]])+
    theme(text = element_text(size = 6),
          axis.title.y = element_blank(),
          panel.background = element_rect(fill = "white"),
          panel.border = element_rect(colour = "black", fill = NA, size = 0.25),
          axis.ticks = element_line(size = 0.25),
          axis.ticks.length = unit(.05, "cm"),
          plot.margin = unit(c(rep(0.75, 4)), "lines"))
}
dens_fig <- ggarrange(plotlist = dens_list, ncol = 4, nrow = 5, labels =
LETTERS[1:length(dens_list)], font.label = list(size = 8))
print(dens_fig)
```

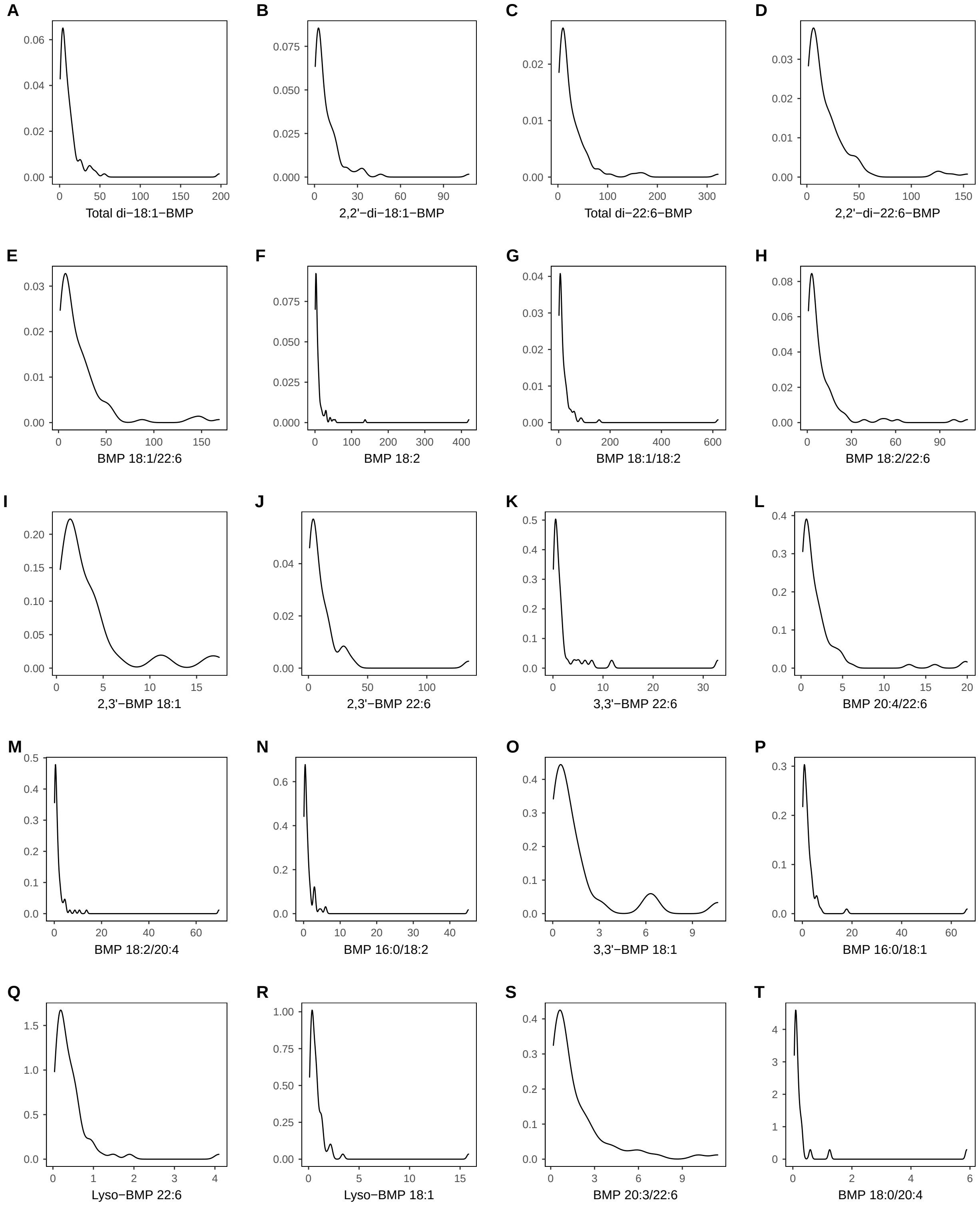

Figure 1: Density plots showing distribution of urine BMP levels data.

```
qq_list = list()
for (i in names(bmp_vars)) {
  qq_list[[i]] = ggplot(df02, aes(sample = !!sym(i))) +
    geom_qq_line(colour = "gray", size = 0.25)+
    geom_qq(shape = "circle", size = 0.25)+
    xlab(bmp_vars[[i]])+
    theme(text = element_text(size = 6),
          axis.title.y = element_blank(),
          panel.background = element_rect(fill = "white"),
          panel.border = element_rect(colour = "black", fill = NA, size = 0.25),
          axis.ticks = element_line(size = 0.25),
          axis.ticks.length = unit(.05, "cm"),
          legend.position = "none",
          plot.margin = unit(c(rep(0.75, 4)), "lines"))
}
qq_fig <- ggarrange(plotlist = qq_list, ncol = 4, nrow = 5, labels =
LETTERS[1:length(qq_list)], font.label = list(size = 8))
print(qq_fig)
```

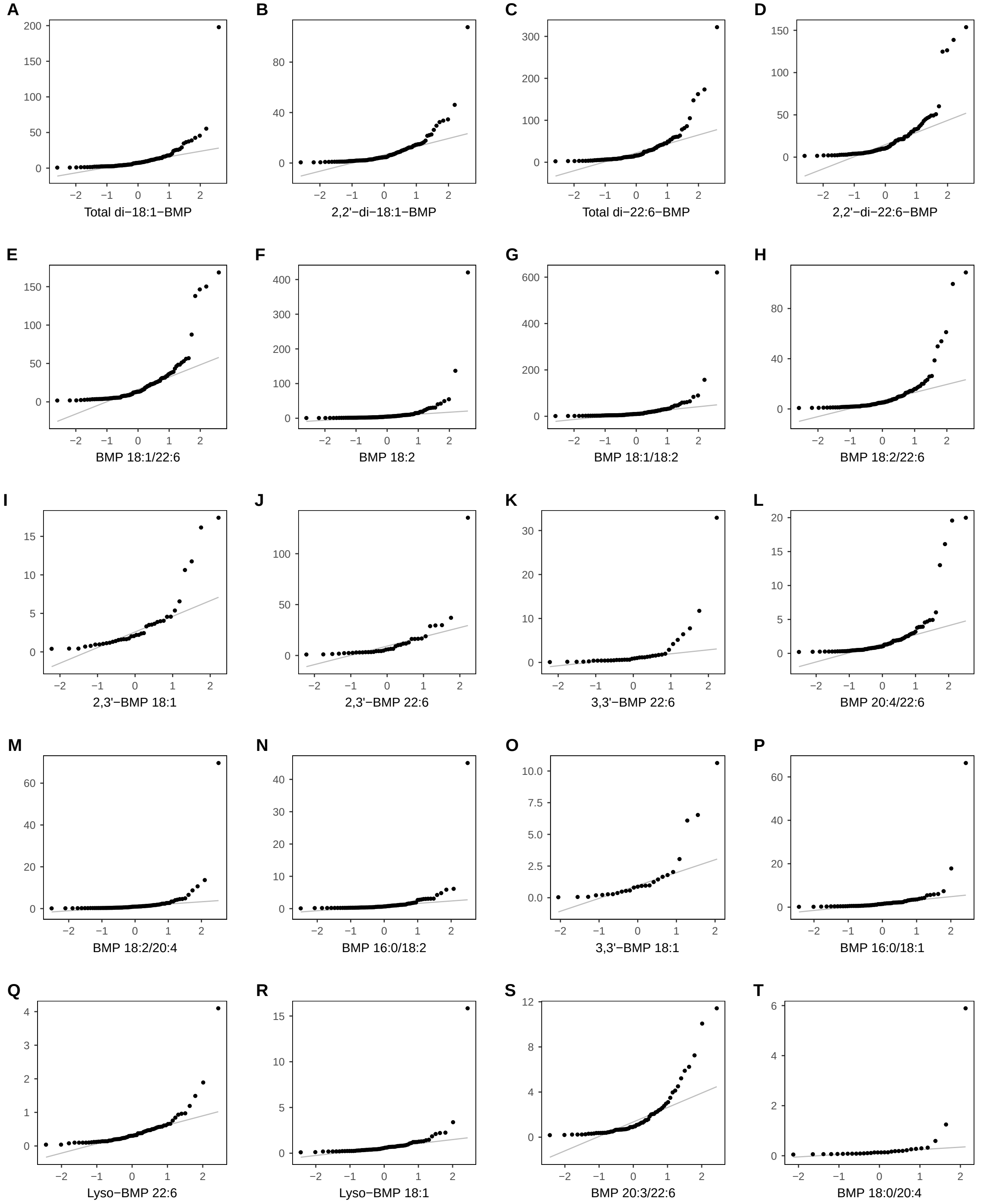

Figure 2: Q-Q plots showing distribution of urine BMP levels data.

#### 2.2 Descriptive statistics

**Table 1** and **Table 2** summarise descriptive statistics for the 4 main BMP species as well as the extended panel, respectively. As the variables studied are not normally distributed, median and range were selected as measures of centre and spread. Overall p values are also reported.

```
# This code chunk uses the raw dataset, not provided.
df01.1 <- subset(df01, group != "Other (SNCA duplication)" & group != "Other (ATP13A2)")
df01.1$group <- factor(df01.1$group, levels = c("Control", "iPD", "LRRK2 G2019S",
"LRRK2 R1441G/C", "VPS35 D620N", "GBA"))

tab01 <- df01.1 %>% mutate(total = 1)%>%
  tbl_summary(include = c(total, age_at_study_participation, sex, disease_duration,
  group, total_di_18_1_bmp, x2_2_di_18_1_bmp, total_di_22_6_bmp, x2_2_di_22_6_bmp),
              missing = "no",
              by = group,
              statistic = list(total ~ "N = {N}",
                               age_at_study_participation ~ "{median} ({min}, {max})",
                               sex ~ "{n}({p}%)",
                               disease_duration ~ "{median} ({min}, {max})",
                               total_di_18_1_bmp ~ "{median} ({min}, {max})",
                               x2_2_di_18_1_bmp ~ "{median} ({min}, {max})",
                               total_di_22_6_bmp ~ "{median} ({min}, {max})",
                               x2_2_di_22_6_bmp ~ "{median} ({min}, {max})"),
              digits = list(total ~ 0,
                            age_at_study_participation ~ 0,
                            sex ~ 0,
                            disease_duration ~ 0,
                            total_di_18_1_bmp ~ 2,
                            x2_2_di_18_1_bmp ~ 2,
                            total_di_22_6_bmp ~ 2,
                            x2_2_di_22_6_bmp ~ 2),
              label = list(total ~ "",
                           age_at_study_participation ~ "Age",
                           sex ~ "Sex",
                           disease_duration ~ "Disease Duration",
                           total_di_18_1_bmp ~ "Total di-18:1-BMP, ng/mg creatinine",
                           x2_2_di_18_1_bmp ~ "2,2'-di-18:1-BMP, ng/mg creatinine",
                           total_di_22_6_bmp ~ "Total 22:6-BMP, ng/mg creatinine",
                          x2_2_di_22_6_bmp ~ "2,2'-di-22:6-BMP, ng/mg creatinine")) %>%
  add_p(include = c(age_at_study_participation,
                    sex,
                    disease_duration,
                    total_di_18_1_bmp,
                    x2_2_di_18_1_bmp,
                    total_di_22_6_bmp,
                    x2_2_di_22_6_bmp))%>%
  modify_header(label = "",
                all_stat_cols() ~ "{level}",
                starts_with("p.value") ~ "p-value") %>%
  as_hux_table()%>%
  huxtable::set_width(1)%>%
  huxtable::set_font_size(6)%>%
  huxtable::set_label("tab:tab01")%>%
  huxtable::set_caption("Summary of descriptive statistics for 4 main BMP species.")%>%
  huxtable::set_top_padding(2)%>%
  huxtable::set_bottom_padding(2)%>%
```

```
  huxtable::set_right_padding(2)%>%
  huxtable::set_left_padding(2)%>%
  huxtable::set_valign("middle")%>%
  huxtable::set_header_rows(1:2, TRUE)%>%
  huxtable::set_header_cols(1, TRUE)%>%
  huxtable::style_headers(bold = TRUE)%>%
  huxtable::set_number_format(1, everywhere, 0)

tab01
```

Table 1: Summary of descriptive statistics for 4 main BMP species.

| | **Control** | **iPD** | **LRRK2 G2019S** | **LRRK2 R1441G/C** | **VPS35 D620N** | **GBA** | **p-value** |
|---|---|---|---|---|---|---|---|
| | **N = 22** | **N = 31** | **N = 18** | **N = 13** | **N = 10** | **N = 10** | |
| **Age** | 54 (24, 71) | 65 (37, 88) | 60 (42, 81) | 65 (50, 83) | 62 (29, 76) | 61 (49, 64) | <0.001 |
| **Sex** | | | | | | | 0.5 |
| **M** | 11(50%) | 16(52%) | 7(39%) | 7(54%) | 3(30%) | 7(70%) | |
| **F** | 11(50%) | 15(48%) | 11(61%) | 6(46%) | 7(70%) | 3(30%) | |
| **Disease Duration** | NA (Inf, -Inf) | 6 (1, 17) | 6 (1, 22) | 9 (6, 20) | 15 (1, 21) | 5 (1, 21) | 0.2 |
| **Total di-18:1-BMP, ng/mg creatinine** | 3.66 (0.72, 8.54) | 6.48 (1.48, 38.88) | 13.43 (3.30, 42.56) | 9.57 (2.70, 34.68) | 15.97 (6.71, 55.37) | 2.56 (0.80, 45.53) | <0.001 |
| **2,2'-di-18:1-BMP, ng/mg creatinine** | 2.04 (0.52, 5.60) | 4.58 (1.01, 34.58) | 11.59 (2.86, 33.59) | 4.68 (1.55, 29.59) | 13.69 (4.54, 46.10) | 1.58 (0.59, 21.59) | <0.001 |
| **Total 22:6-BMP, ng/mg creatinine** | 8.33 (2.18, 41.14) | 12.84 (3.28, 104.92) | 43.52 (6.98, 321.98) | 28.11 (7.93, 81.34) | 29.19 (15.51, 162.36) | 9.66 (3.98, 34.21) | <0.001 |
| **2,2'-di-22:6-BMP, ng/mg creatinine** | 4.83 (1.47, 19.59) | 9.86 (2.17, 60.15) | 33.43 (5.61, 153.74) | 21.27 (4.35, 47.35) | 22.61 (8.98, 126.35) | 5.93 (2.81, 21.31) | <0.001 |

**N = N; Median (Range); n(%)**

**Kruskal-Wallis rank sum test; Fisher's exact test**

```
tab01 %>% huxtable::as_flextable() %>% flextable::save_as_docx(path =
here("table01.docx"))
```

```
df02.1 <- subset(df02, group != "Other (SNCA duplication)" & group != "Other (ATP13A2)")
df02.1$group <- factor(df02.1$group, levels = c("Control", "iPD", "LRRK2 G2019S",
"LRRK2 R1441G/C", "VPS35 D620N", "GBA"))

temp_vars <- bmp_vars
temp_vars$total <- ""

supp_tab02 <- df02.1 %>% mutate(total = 1) %>%
  tbl_summary(by = group,
              missing = "no",
              include = c(total, names(bmp_vars[c(5:20)])),
              statistic = list(all_continuous() ~ "{median} ({min}, {max})",
                               total ~ "N = {n}"),
              digits = list(all_continuous() ~ 2,
                            total ~ 0),
              label = temp_vars[5:21]) %>%
  add_p(include = c(names(temp_vars[c(5:20)])))%>%
  modify_header(label = "",
                all_stat_cols() ~ "{level}",
                starts_with("p.value") ~ "p-value") %>%
  as_hux_table()%>%
```

```
  huxtable::set_width(1)%>%
  huxtable::set_font_size(6)%>%
  huxtable::set_label("tab:stab02")%>%
  huxtable::set_caption("Summary of descriptive statistics for extended panel of BMP
species.")%>%
  huxtable::set_top_padding(2)%>%
  huxtable::set_bottom_padding(2)%>%
  huxtable::set_right_padding(2)%>%
  huxtable::set_left_padding(2)%>%
  huxtable::set_valign("middle")%>%
  huxtable::set_header_rows(1:2, TRUE)%>%
  huxtable::set_header_cols(1, TRUE)%>%
  huxtable::style_headers(bold = TRUE)%>%
  huxtable::set_number_format(1, everywhere, 0)

supp_tab02
```

Table 2: Summary of descriptive statistics for extended panel of BMP species.

| | **Control** | **iPD** | **LRRK2 G2019S** | **LRRK2 R1441G/C** | **VPS35 D620N** | **GBA** | **p-value** |
|---|---|---|---|---|---|---|---|
| | **N = 22** | **N = 31** | **N = 18** | **N = 13** | **N = 10** | **N = 10** | |
| **BMP 18:1/22:6** | 5.19 (1.86, 22.99) | 10.38 (2.92, 56.06) | 32.40 (12.75, 168.50) | 19.42 (5.80, 53.00) | 25.37 (8.59, 146.41) | 3.87 (1.74, 48.30) | <0.001 |
| **BMP 18:2** | 2.02 (0.72, 6.25) | 2.97 (0.95, 29.79) | 9.88 (2.08, 136.82) | 9.01 (2.20, 27.02) | 16.67 (5.98, 42.34) | 1.97 (0.95, 9.82) | <0.001 |
| **BMP 18:1/18:2** | 4.57 (1.14, 15.29) | 8.07 (2.16, 58.99) | 23.98 (5.66, 157.33) | 18.60 (4.40, 46.40) | 28.19 (9.83, 89.80) | 3.48 (1.63, 53.08) | <0.001 |
| **BMP 18:2/22:6** | 1.94 (0.77, 7.56) | 3.51 (1.25, 18.31) | 13.70 (4.72, 108.74) | 8.08 (4.13, 26.27) | 12.98 (5.17, 53.91) | 1.92 (0.89, 8.00) | <0.001 |
| **2,3'-BMP 18:1** | 1.43 (0.43, 4.04) | 3.88 (0.78, 11.75) | 9.73 (3.31, 16.15) | 2.21 (1.41, 6.56) | NA (Inf, -Inf) | 1.39 (0.40, 17.42) | 0.074 |
| **2,3'-BMP 22:6** | 3.81 (0.96, 16.42) | 10.15 (2.27, 37.03) | 82.42 (29.50, 135.33) | 16.22 (3.27, 29.80) | NA (Inf, -Inf) | 3.94 (1.78, 11.63) | 0.013 |
| **3,3'-BMP 22:6** | 0.60 (0.08, 5.13) | 0.84 (0.17, 7.75) | 22.32 (11.73, 32.91) | 1.70 (0.42, 4.19) | NA (Inf, -Inf) | 0.53 (0.15, 1.27) | 0.019 |
| **BMP 20:4/22:6** | 0.48 (0.27, 1.94) | 0.65 (0.21, 4.93) | 2.13 (0.53, 19.57) | 1.61 (0.52, 6.04) | 2.66 (1.31, 16.10) | 0.70 (0.23, 2.21) | <0.001 |
| **BMP 18:2/20:4** | 0.31 (0.15, 1.40) | 0.47 (0.19, 4.03) | 1.31 (0.44, 13.64) | 1.32 (0.27, 4.53) | 2.50 (1.02, 10.64) | 0.72 (0.11, 2.09) | <0.001 |
| **BMP 16:0/18:2** | 0.30 (0.19, 0.82) | 0.36 (0.09, 4.23) | 1.07 (0.35, 6.13) | 1.00 (0.42, 3.06) | 1.72 (0.58, 5.87) | 0.67 (0.17, 3.10) | <0.001 |
| **3,3'-BMP 18:1** | 0.37 (0.04, 1.79) | 1.09 (0.80, 6.09) | 5.43 (0.22, 10.63) | 1.65 (0.19, 3.05) | NA (Inf, -Inf) | 0.48 (0.27, 6.53) | 0.2 |
| **BMP 16:0/18:1** | 0.71 (0.17, 1.02) | 0.71 (0.15, 3.87) | 2.19 (0.55, 6.06) | 1.96 (0.81, 5.44) | 3.64 (1.38, 17.82) | 0.41 (0.30, 2.36) | <0.001 |
| **Lyso-BMP 22:6** | 0.11 (0.04, 0.30) | 0.18 (0.10, 0.93) | 0.50 (0.13, 1.49) | 0.41 (0.20, 0.61) | 0.52 (0.21, 1.89) | 0.39 (0.10, 0.66) | <0.001 |
| **Lyso-BMP 18:1** | 0.25 (0.20, 0.71) | 0.38 (0.12, 2.10) | 0.75 (0.25, 1.85) | 0.76 (0.39, 1.29) | 0.83 (0.42, 3.39) | 0.59 (0.10, 2.21) | 0.001 |
| **BMP 20:3/22:6** | 0.35 (0.20, 0.45) | 0.69 (0.23, 3.11) | 2.03 (0.67, 10.07) | 1.24 (0.66, 2.60) | 2.62 (1.29, 11.42) | 0.45 (0.18, 0.72) | <0.001 |
| **BMP 18:0/20:4** | 0.07 (0.07, 0.08) | 0.07 (0.05, 0.19) | 0.12 (0.08, 0.59) | NA (Inf, -Inf) | 0.19 (0.09, 1.25) | 0.28 (0.28, 0.28) | 0.015 |

**N = n; Median (Range)**

**Kruskal-Wallis rank sum test**

```
supp_tab02 %>% huxtable::as_flextable() %>% flextable::save_as_docx(path =
here("supp_table02.docx"))
```

#### 2.3 Data visualisation

BMP variation per experimental group was visualised by plotting boxplots for each BMP species (**Figure 3**). In cases where overall $p < 0.05$ and at least 3 datapoints were available per group, comparisons to the `Control` group, calculated by Dunn's post-hoc test, are shown in the plot. Complete multiple comparisons for all possible pairwise comparisons are summarised in **Section 3.1**.

```
makeboxplot = function(dataset,    # When calling function, use dataset name as object
                      x_var,       # Provide all other arguments as strings
                      y_var,
```

```
                    fill_var,
                    shape_var,
                    y_lab,
                    ref_group) {

  # Get values from variable names
  x <- eval(parse(text=paste0(deparse(substitute(dataset)),"$",x_var)))
  y <- eval(parse(text=paste0(deparse(substitute(dataset)),"$",y_var)))
  fill <- eval(parse(text=paste0(deparse(substitute(dataset)),"$",fill_var)))

  # Set colour palette to use in plots, based on number of levels of factor to plot by.
  lab_col <- pal_locuszoom("default")(nlevels(x))

  # Save N number per group on vector named counts. This will be used to add N to the
  plot.
  counts = dataset %>% group_by(!!sym(x_var)) %>% tally

  # Make statistics table with p-values and positions to plot on. Use Kruskal-Wallis
  for     overall p value and Dunn's test for multiple comparisons.
  formula <- paste(y_var, "~", fill_var)
  stats <- kruskal.test(formula=as.formula(formula), data=dataset)
  pval <- stats$p.value
  dunn <- dunn_test(data = dataset, formula = as.formula(formula), p.adjust.method =
"fdr") %>%   add_xy_position(x = x_var, fun = "max", step.increase = 0.05) %>%
subset(group1 == ref_group)

  # Make formatted boxplot.
  boxp <- ggplot(dataset, aes(x = !!sym(x_var),
                              y = !!sym(y_var)))+
    geom_boxplot(aes(fill = !!sym(fill_var)), outlier.shape = NA, lwd = 0.25)+
    scale_fill_locuszoom(drop = FALSE)+
    scale_x_discrete(labels = function(x) str_wrap(x, width = 10))+
    theme_light(base_size = 3.5)+
    theme(plot.title = element_blank(),
          axis.title.x = element_blank(),
          axis.text.x = element_text(face="bold"),
          axis.title.y = element_text(face = "bold"),
          legend.position = "none")+
    guides(fill="none")+
    ylab(y_lab)+
    geom_jitter((aes(shape = !!sym(shape_var), fill = !!sym(fill_var))),
                width = 0.3,
                alpha = 0.5,
                stroke = 0.1,
                size = 1)+
    scale_shape_manual(values=c(25, 21, 21))+
    geom_text(data=counts, aes(label=paste("n=",n,sep = ""),
                               y=min(y)-0.08*max(y),
                               fontface = "bold"),
              size = 1.5,
              position=position_dodge(0.9),
              angle = 0,
              colour = lab_col)

  # calculations for yposition of stat brackets distribution.
  bars <- nlevels(x) - 2
```

```
  max_plot <- max(layer_data(boxp)$ymax)
  top_vec <- rep(max_plot, times = (bars+1))
  mult_vec <- c(0:bars)
  step_increase = (max(y, na.rm = TRUE)-max_plot)/(bars+1)

  mult_vec <- mult_vec * step_increase
  top_vec <- top_vec + mult_vec
  dunn$y.position <- top_vec

  # add p values/brackets to plot
  if(pval >= 0.05 || any(counts$n < 3)) {
    boxp <- boxp +
      stat_compare_means(label.y = max(y, na.rm = TRUE),
                         label.x = 1.3,
                         size = 1.5)
  } else {
    boxp <- boxp +
      stat_compare_means(label.y = max(y, na.rm = TRUE),
                         label.x = 1.3,
                         size = 1.5)+
      stat_pvalue_manual(dunn, label = 'p.adj.signif', tip.length = 0.01, size = 1.5)
  }}
```

```
group_var <- "group"
shape_var <- "pd_status"
ref_group <- "Control"
units <- "\n (ng/mg creatinine)"

boxp_list = list()
for(i in names(bmp_vars)) {
  tempdf <- drop_na(df02.1, !!sym(i))
  tempdf <- droplevels(tempdf)
  boxp_list[[i]] = makeboxplot(dataset = tempdf,
                               x_var = group_var,
                               y_var = names(bmp_vars[i]),
                               fill_var = group_var,
                               shape_var = "pd_status",
                               y_lab = paste0(bmp_vars[[i]], units),
                               ref_group = ref_group)}

boxp_fig <- ggarrange(plotlist = boxp_list, ncol = 4, nrow = 5, labels =
LETTERS[1:length(qq_list)], font.label = list(size = 8))
print(boxp_fig)
```

```
# Uncomment below to export
# fig01 <- ggarrange(plotlist = boxp_list[1:4], ncol = 2, nrow = 2, labels =
LETTERS[1:length(qq_list)], font.label = list(size = 4))
# ggsave(filename = here::here("figure01.pdf"), plot = fig01, width = 5, height = 4,
units = "in")

# figS1 <- garrange(plotlist = boxp_list[5:20], ncol = 2, nrow = 8, labels =
LETTERS[1:length(qq_list)], font.label = list(size = 8))
# ggsave(filename = here::here("figureS1.pdf"), plot = fig02,  width = 6, height = 5,
units = "in")
```

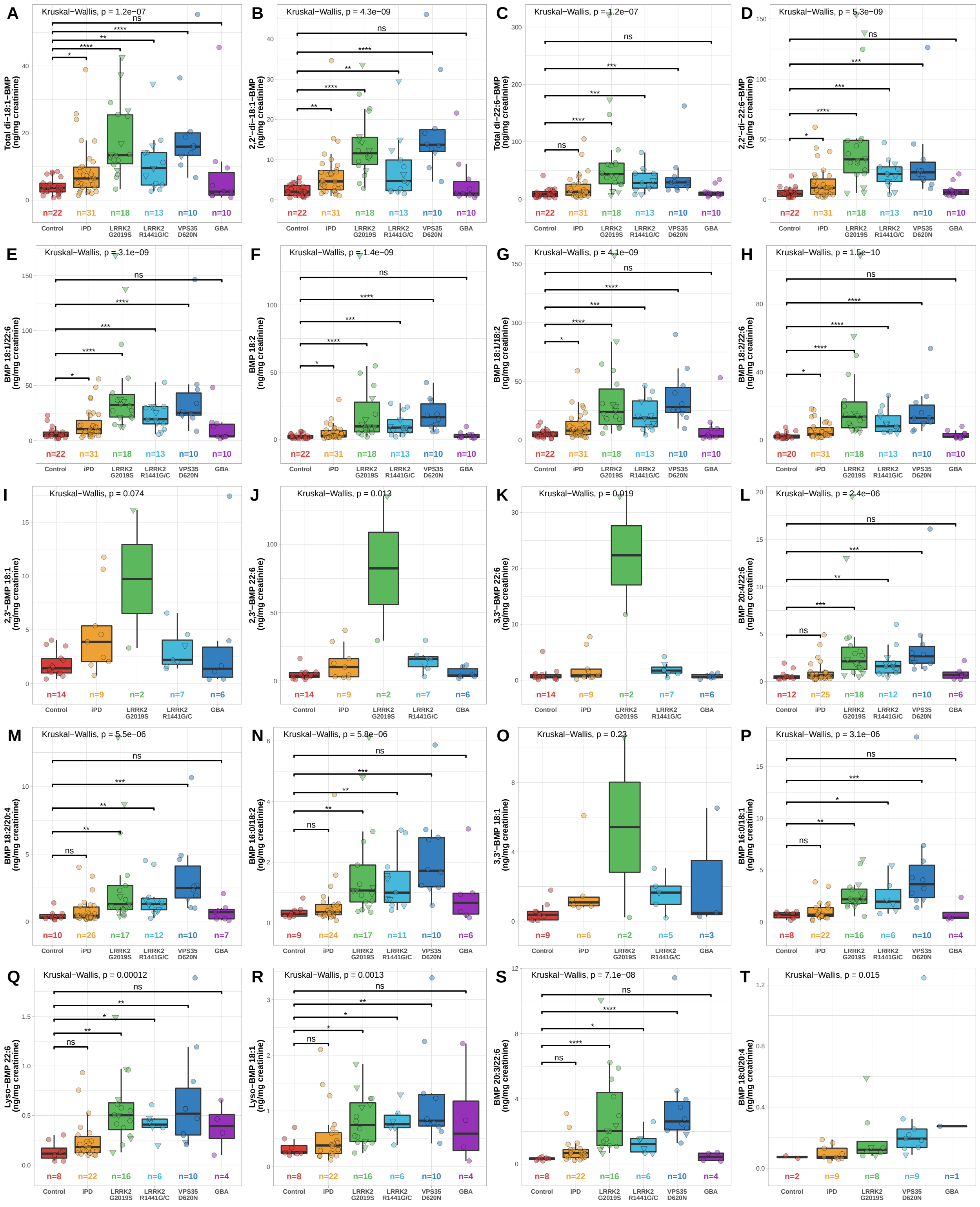

Figure 3: Boxplots showing BMP data per experimental group.

#### 3 Statistical analysis

##### 3.1 Urine BMP variation per group

In **Section 2.3**, statistically significant differences to the Control group are displayed in boxplots. Here, all possible pairwise comparisons are computed by Dunn's test and summarised in **Supplementary Table 3**. Multiple comparisons are only computed and displayed only in cases where Kruskal-Wallis test returns overall $p < 0.05$, and at least 3 datapoints are available per compared group.

```
suppl_tabl_03 <- data.frame()
col_lab <- c("BMP", "Group 1", "Group 2", "N1", "N2", "Statistic", "p", "p adjusted")
for(i in names(bmp_vars)) {
  tempdf <- drop_na(df02.1, !!sym(i))
  tempdf <- droplevels(tempdf)
  counts = tempdf %>% group_by(group) %>% tally
  stat_formula <- paste(names(bmp_vars[i]), "~ group")
  stats <- kruskal.test(formula = as.formula(stat_formula), data=tempdf)
  pval <- stats$p.value
  if(pval < 0.05 && all(counts$n >= 3)) {
    dunn <- dunn_test(data = tempdf,
                      formula=as.formula(stat_formula),
                      p.adjust.method = "BH")
    suppl_tabl_03 <- rbind(suppl_tabl_03, dunn)
  }}

for(i in names(bmp_vars)) {
  suppl_tabl_03[suppl_tabl_03 == i] <- bmp_vars[[i]]
}

colnames(suppl_tabl_03) <- col_lab
write_csv(suppl_tabl_03, here::here("suppl_tabl_03.csv"))
```

##### 3.2 Urine BMP variation in PD vs NMC

Kruskal-Wallis tests were carried out to compare PD versus NMC for each BMP species. In all cases, either $p > 0.05$ or there were less than 3 datapoints in one particular group, so no post-hoc tests were applicable.

Kruskal-Wallis p values are summarised in **Supplementary Table 4**. In the case of BMP 20:3/22:6, although $p < 0.05$, there was only 1 datapoint in the VPS35 D620N - NMC group, hence no multiple comparison analysis was carried out.

```
selected <- c("LRRK2 G2019S", "LRRK2 R1441G/C", "VPS35 D620N")
df02.2 <- df02.1[df02.1$group %in% selected, ]

non_significant <- c()
suppl_tabl_02.1 <- data.frame()
suppl_tabl_02.2 <- data.frame(matrix(ncol = 7, nrow = 0))
col_lab <-  c("BMP", "Group 1", "Group 2", "N1", "N2", "Statistic", "p-value")

for(i in names(bmp_vars)) {
  tempdf <- drop_na(df02.2, !!sym(i))
  tempdf <- droplevels(tempdf)
  counts = tempdf %>% group_by(group, pd_status) %>% tally
  stat_formula <- paste(names(bmp_vars[i]), "~ pd_status")
  stats <- kruskal.test(formula = as.formula(stat_formula), data=tempdf)
  pval <- stats$p.value
  if(pval < 0.05 && all(counts$n > 3)) {
```

```
    dunn <- tempdf %>% group_by(group) %>% dunn_test(formula=as.formula(stat_formula),
                                                     p.adjust.method = "BH")
    suppl_tabl_02.1 <- rbind(suppl_tabl_02.1, dunn) #returns empty table as no values
return TRUE on if statement
  }
  else(
    non_significant <- append(non_significant, names(bmp_vars[i]))
  )}

for(i in non_significant){
  tempdf <- drop_na(df02.2, !!sym(i))
  tempdf <- droplevels(tempdf)
  counts = tempdf %>% group_by(pd_status) %>% tally
  stat_formula <- paste(names(bmp_vars[i]), "~ pd_status")
  stats <- kruskal.test(formula = as.formula(stat_formula), data=tempdf)
  sval <- stats$statistic
  pval <- stats$p.value
  new_row <- c(bmp_vars[[i]], "PD", "NMC", counts$n[2], counts$n[1], sval, pval)
  suppl_tabl_02.2 <- rbind(suppl_tabl_02.2, new_row)
}

colnames(suppl_tabl_02.2) <- col_lab
write_csv(suppl_tabl_02.2, here::here("suppl_tabl_02.csv"))
```

#### 3.3 Effect of covariates on urine BMP levels

To simultaneously interrogate the effect of different covariates on urine BMP levels, for the 4 main BMP species, linear models were computed. Since the BMP data is not normally distributed, the data was first log-transformed. Density plots and q-q plots in **Figure 4** show that log-transformed data is normally distributed and can be used as response variables for linear models. Note that, in this section, the raw dataset including participants' ages (not provided) was used.

```
# New dataset with log-transformed BMP values
df01.2 <- df01.1
df01.2[bmp_variables] <- log(df01.2[bmp_variables])

# Check for normal distribution of 4 main BMP species
dens_list = list()
for (i in names(bmp_vars[1:4])) {
  dens_list[[i]] = ggplot(df01.2, aes(x = !!sym(i))) +
    geom_density(alpha = 0.6,
                 size = 0.25)+
    xlab(bmp_vars[[i]])+
    theme(text = element_text(size = 6),
          axis.title.y = element_blank(),
          panel.background = element_rect(fill = "white"),
          panel.border = element_rect(colour = "black", fill = NA, size = 0.25),
          axis.ticks = element_line(size = 0.25),
          axis.ticks.length = unit(.05, "cm"))
}

qq_list = list()
for (i in names(bmp_vars[1:4])) {
  qq_list[[i]] = ggplot(df01.2, aes(sample = !!sym(i))) +
    geom_qq_line(colour = "gray", size = 0.25)+
```

```
    geom_qq(shape = "circle", size = 0.25)+
    xlab(bmp_vars[[i]])+
    theme(text = element_text(size = 6),
          axis.title.y = element_blank(),
          panel.background = element_rect(fill = "white"),
          panel.border = element_rect(colour = "black", fill = NA, size = 0.25),
          axis.ticks = element_line(size = 0.25),
          axis.ticks.length = unit(.05, "cm"),
          legend.position = "none")
}

fig_list <- c(rbind(dens_list,qq_list))

fig <- ggarrange(plotlist = fig_list, ncol = 4, nrow = 2, labels =
LETTERS[1:length(fig_list)], font.label = list(size = 6))
print(fig)
```

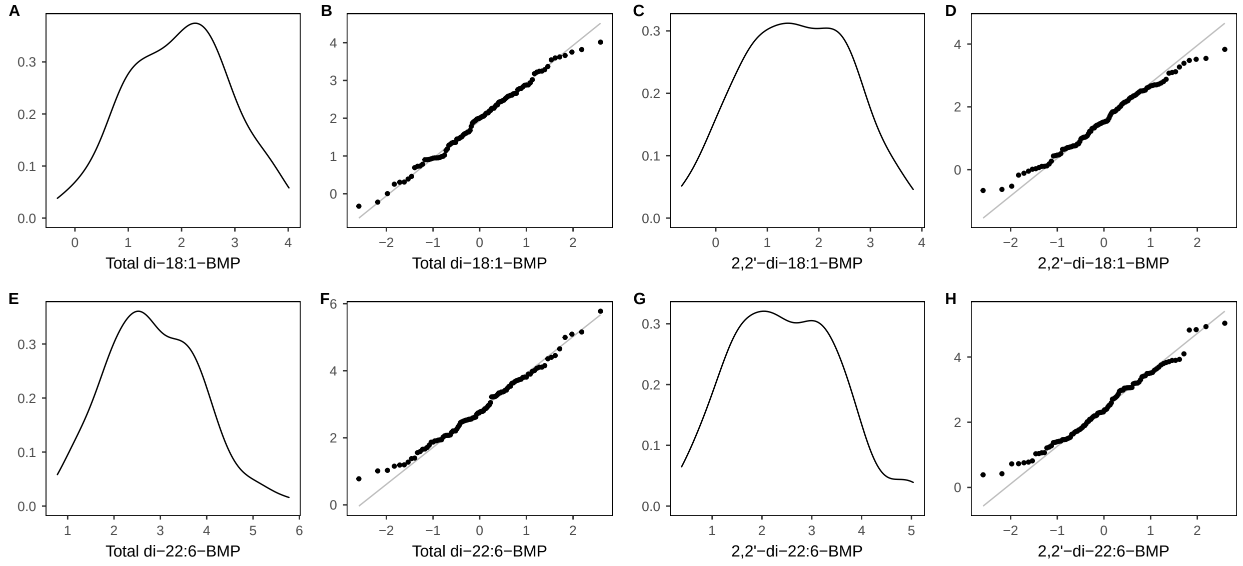

Figure 4: Density plots and q-q plots for log-transformed data of 4 main BMP species shows data is now normally distibuted.

In each subsection below, a the maximal model was generated taking one of the 4 main BMP species as a response variable and group, age, and sex as covariates. This was reduced to a minimum sufficient model by sequential removal of non-significant terms. The behaviour of residuals was checked by plotting and the models were considered valid only if residuals showed constant variation across the range as well as normal distribution.

##### 3.3.1 *Total di-18:1-BMP*

The minimum sufficient model **(1)** included only group as a predictor **(Table 3)**. Statistically significant effects are observed for LRRK2 and VPS35 mutation carriers, consistently with results reported in **Section 3**.

**Figure 5** shows residuals are evenly distributed across range, and follow a normal distribution (q-q plot), confirming the validity of the model.

```
m1 <- lm(total_di_18_1_bmp~group+age_at_study_participation+sex, data=df01.2)
m1.1 <- lm(total_di_18_1_bmp~group, data=df01.2)
models <- list(m1, m1.1)
tab03 <- modelsummary(models,
             output = "huxtable",
             statistic = "p.value",
             stars = c("*" = .05, "**" = .01, "***" = .001),
             shape = term ~ model + statistic,
             sparse_header = TRUE,
             coef_rename = c("groupiPD" = "iPD",
                             "groupLRRK2 G2019S" = "LRRK2 G2019S",
                             "groupLRRK2 R1441G/C" = "LRRK2 R1441G/C",
                             "groupVPS35 D620N" = "VPS35 D620N",
                             "groupGBA" = "GBA",
                             "sexF" = "Sex",
                             "age_at_study_participation" = "Age"))

column_headers <- c("", "Model 1", "", "Model 2", "")
tab03 <-rbind(column_headers, tab03)
tab03[2,2] <- "Estimate"
tab03[2,3] <- "p-value"
tab03[2,4] <- "Estimate"
tab03[2,5] <- "p-value"

tab03 %>%
  huxtable::set_font_size(7)%>%
  huxtable::set_label("tab:tab03")%>%
  huxtable::set_caption("Summary of maximal model (1) and minimum sufficient model (2)
for total di-18:1-BMP.")%>%
  huxtable::set_top_padding(2)%>%
  huxtable::set_bottom_padding(2)%>%
  huxtable::set_right_padding(2)%>%
  huxtable::set_left_padding(2)%>%
  huxtable::set_valign("middle")%>%
  huxtable::set_header_rows(1:2, TRUE)%>%
  huxtable::set_header_cols(1, TRUE)%>%
  huxtable::style_headers(bold = TRUE)%>%
  huxtable::merge_cells(1, 2:3)%>%
  huxtable::merge_cells(1, 4:5)%>%
  huxtable::set_align(everywhere, 2:5, "center")
```

```
par(mfrow = c(1,4))
plot(m1.1)
```

Table 3: Summary of maximal model (1) and minimum sufficient model (2) for total di-18:1-BMP.

| | Model 1 | | Model 2 | |
|---|---|---|---|---|
| | Estimate | p-value | Estimate | p-value |
| **(Intercept)** | 0.696 | 0.065 | 1.187*** | 0.000 |
| **iPD** | 0.474 | 0.051 | 0.678** | 0.003 |
| **LRRK2 G2019S** | 1.339*** | 0.000 | 1.496*** | 0.000 |
| **LRRK2 R1441G/C** | 0.716* | 0.018 | 0.960*** | 0.001 |
| **VPS35 D620N** | 1.581*** | 0.000 | 1.671*** | 0.000 |
| **GBA** | -0.066 | 0.828 | 0.108 | 0.718 |
| **Age** | 0.013 | 0.067 | | |
| **Sex** | -0.242 | 0.121 | | |
| **Num.Obs.** | 104 | | 104 | |
| **R2** | 0.407 | | 0.373 | |
| **R2 Adj.** | 0.363 | | 0.341 | |
| **AIC** | 250.4 | | 252.2 | |
| **BIC** | 274.2 | | 270.7 | |
| **Log.Lik.** | -116.216 | | -119.109 | |
| **RMSE** | 0.74 | | 0.76 | |

*** p $<$ 0.05, ** p $<$ 0.01, *** p $<$ 0.001**

###### 3.3.2 *2,2'-di-18:1-BMP*

The minimum sufficient model **(1)** included only group as a predictor **(Table 4)**. Statistically significant effects are observed for LRRK2 and VPS35 mutation carriers, consistently with results reported in **Section 3**.

**Figure 6** shows residuals are evenly distributed across range, and follow a normal distribution (q-q plot), confirming the validity of the model.

```
m2 <- lm(x2_2_di_18_1_bmp~group+age_at_study_participation+sex, data=df01.2)
m2.1 <- lm(x2_2_di_18_1_bmp~group, data=df01.2)
models <- list(m2, m2.1)
tab04 <- modelsummary(models,
             output = "huxtable",
```

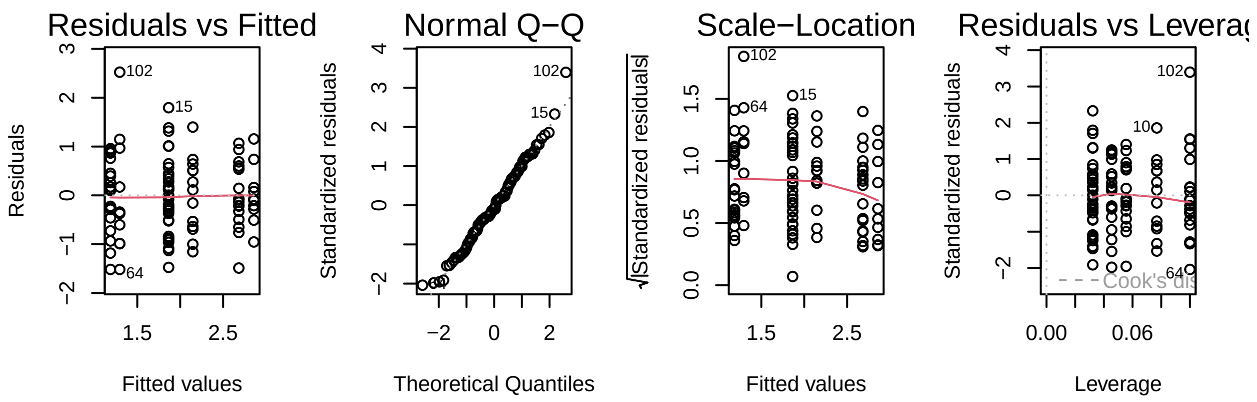

Figure 5: Residuals behaviour for the minimum sufficient model of total di-18:1-BMP.

```
                 statistic = "p.value",
                 stars = c("*" = .05, "**" = .01, "***" = .001),
                 shape = term ~ model + statistic,
                 sparse_header = TRUE,
                 coef_rename = c("groupiPD" = "iPD",
                                 "groupLRRK2 G2019S" = "LRRK2 G2019S",
                                 "groupLRRK2 R1441G/C" = "LRRK2 R1441G/C",
                                 "groupVPS35 D620N" = "VPS35 D620N",
                                 "groupGBA" = "GBA",
                                 "sexF" = "Sex",
                                 "age_at_study_participation" = "Age"))

column_headers <- c("", "Model 1", "", "Model 2", "")
tab04 <-rbind(column_headers, tab04)
tab04[2,2] <- "Estimate"
tab04[2,3] <- "p-value"
tab04[2,4] <- "Estimate"
tab04[2,5] <- "p-value"

tab04 %>%
  huxtable::set_font_size(7)%>%
  huxtable::set_label("tab:tab04")%>%
  huxtable::set_caption("Summary of maximal model (1) and minimum sufficient model (2)
for 2,2'-di-18:1-BMP.")%>%
  huxtable::set_top_padding(2)%>%
  huxtable::set_bottom_padding(2)%>%
  huxtable::set_right_padding(2)%>%
  huxtable::set_left_padding(2)%>%
  huxtable::set_valign("middle")%>%
  huxtable::set_header_rows(1:2, TRUE)%>%
  huxtable::set_header_cols(1, TRUE)%>%
  huxtable::style_headers(bold = TRUE)%>%
  huxtable::merge_cells(1, 2:3)%>%
  huxtable::merge_cells(1, 4:5)%>%
  huxtable::set_align(everywhere, 2:5, "center")
```

```
par(mfrow = c(1,4))
plot(m2.1)
```

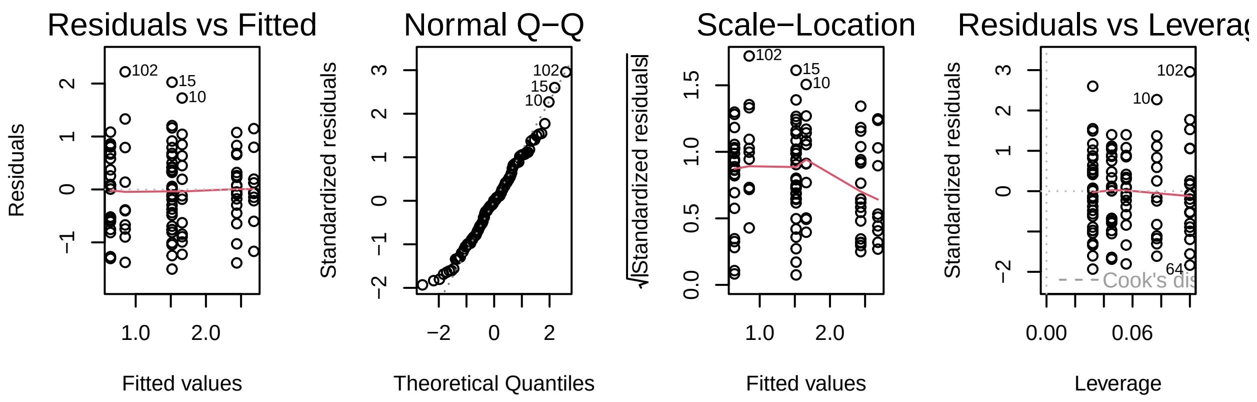

Figure 6: Residuals behaviour for the minimum sufficient model of 2,2'-di-18:1-BMP.

Table 4: Summary of maximal model (1) and minimum sufficient model (2) for 2,2'-di-18:1-BMP.

| | Model 1 | | Model 2 | |
|---|---|---|---|---|
| | Estimate | p-value | Estimate | p-value |
| (Intercept) | 0.195 | 0.606 | 0.641*** | 0.000 |
| iPD | 0.682** | 0.006 | 0.877*** | 0.000 |
| LRRK2 G2019S | 1.652*** | 0.000 | 1.798*** | 0.000 |
| LRRK2 R1441G/C | 0.785* | 0.010 | 1.021*** | 0.000 |
| VPS35 D620N | 1.965*** | 0.000 | 2.041*** | 0.000 |
| GBA | 0.033 | 0.913 | 0.209 | 0.489 |
| Age | 0.012 | 0.082 | | |
| Sex | -0.279 | 0.077 | | |
| Num.Obs. | 104 | | 104 | |
| R2 | 0.482 | | 0.450 | |
| R2 Adj. | 0.444 | | 0.422 | |
| AIC | 252.2 | | 254.4 | |
| BIC | 276.0 | | 272.9 | |
| Log.Lik. | -117.098 | | -120.188 | |
| RMSE | 0.75 | | 0.77 | |
| * $p < 0.05$, ** $p < 0.01$, *** $p < 0.001$ | | | | |

##### 3.3.3 *Total di-22:6-BMP*

The minimum sufficient model **(1)** included both group and age as predictors **(Table 5)**. Even when adjusted for age, statistically significant effects are observed for LRRK2 and VPS35 mutation carriers, consistently with results reported in **Section 3**.

**Figure 7** shows residuals are evenly distributed across range, and follow a normal distribution (q-q plot), confirming the validity of the model.

```
m3 <- lm(total_di_22_6_bmp~group+age_at_study_participation+sex, data=df01.2)
m3.1 <- lm(total_di_22_6_bmp~group+age_at_study_participation, data=df01.2)
models <- list(m3, m3.1)
tab05 <- modelsummary(models,
             output = "huxtable",
             statistic = "p.value",
             stars = c("*" = .05, "**" = .01, "***" = .001),
             shape = term ~ model + statistic,
             sparse_header = TRUE,
             coef_rename = c("groupiPD" = "iPD",
                             "groupLRRK2 G2019S" = "LRRK2 G2019S",
                             "groupLRRK2 R1441G/C" = "LRRK2 R1441G/C",
                             "groupVPS35 D620N" = "VPS35 D620N",
                             "groupGBA" = "GBA",
                             "sexF" = "Sex",
                             "age_at_study_participation" = "Age"))

column_headers <- c("", "Model 1", "", "Model 2", "")
tab05 <-rbind(column_headers, tab05)
tab05[2,2] <- "Estimate"
```

```r
tab05[2,3] <- "p-value"
tab05[2,4] <- "Estimate"
tab05[2,5] <- "p-value"

tab05 %>%
  huxtable::set_font_size(7)%>%
  huxtable::set_label("tab:tab05")%>%
  huxtable::set_caption("Summary of maximal model (1) and minimum sufficient model (2)
for total di-22:6-BMP.")%>%
  huxtable::set_top_padding(2)%>%
  huxtable::set_bottom_padding(2)%>%
  huxtable::set_right_padding(2)%>%
  huxtable::set_left_padding(2)%>%
  huxtable::set_valign("middle")%>%
  huxtable::set_header_rows(1:2, TRUE)%>%
  huxtable::set_header_cols(1, TRUE)%>%
  huxtable::style_headers(bold = TRUE)%>%
  huxtable::merge_cells(1, 2:3)%>%
  huxtable::merge_cells(1, 4:5)%>%
  huxtable::set_align(everywhere, 2:5, "center")
```

Table 5: Summary of maximal model (1) and minimum sufficient model (2) for total di-22:6-BMP.

| | **Model 1** | | **Model 2** | |
|---|---|---|---|---|
| | **Estimate** | **p-value** | **Estimate** | **p-value** |
| **(Intercept)** | 0.651 | 0.076 | 0.661 | 0.068 |
| **iPD** | 0.150 | 0.525 | 0.148 | 0.527 |
| **LRRK2 G2019S** | 1.349*** | 0.000 | 1.351*** | 0.000 |
| **LRRK2 R1441G/C** | 0.733* | 0.013 | 0.731* | 0.013 |
| **VPS35 D620N** | 1.086*** | 0.000 | 1.091*** | 0.000 |
| **GBA** | 0.016 | 0.957 | 0.009 | 0.975 |
| **Age** | 0.029*** | 0.000 | 0.029*** | 0.000 |
| **Sex** | 0.029 | 0.850 | | |
| **Num.Obs.** | 104 | | 104 | |
| **R2** | 0.489 | | 0.488 | |
| **R2 Adj.** | 0.451 | | 0.457 | |
| **AIC** | 245.0 | | 243.1 | |
| **BIC** | 268.8 | | 264.2 | |
| **Log.Lik.** | -113.510 | | -113.530 | |
| **RMSE** | 0.72 | | 0.72 | |
| *** p < 0.05, ** p < 0.01, *** p < 0.001** | | | | |

```r
par(mfrow = c(1,4))
plot(m3.1)
```

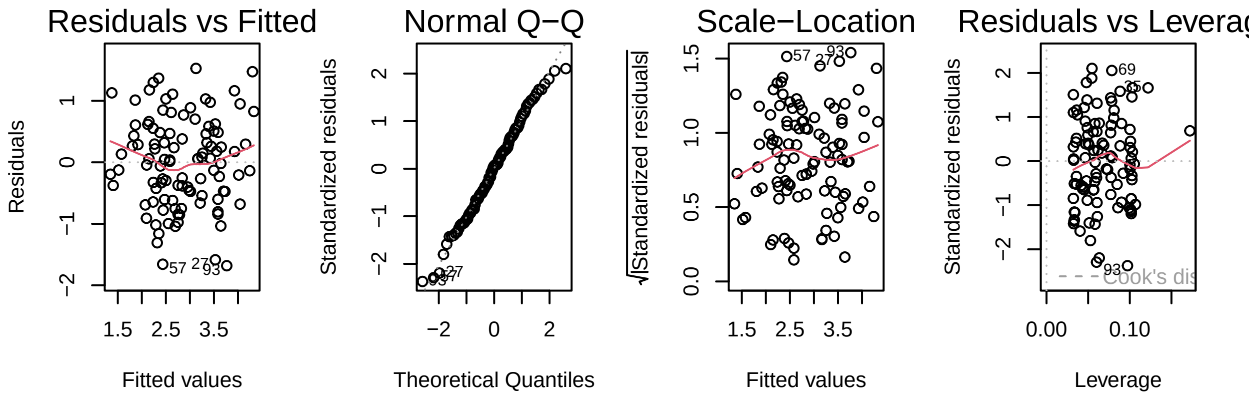

Figure 7: Residuals behaviour for the minimum sufficient model of total di-22:6-BMP.

###### 3.3.4 *2,2'-di-22:6-BMP*

The minimum sufficient model **(1)** included both group and age as predictors **(Table 6)**. Even when adjusted for age, statistically significant effects are observed for LRRK2 and VPS35 mutation carriers, consistently with results reported in **Section 3**.

**Figure 8** shows residuals are evenly distributed across range, and follow a normal distribution (q-q plot), confirming the validity of the model.

```
m4 <- lm(x2_2_di_22_6_bmp~group+age_at_study_participation+sex, data=df01.2)
m4.1 <- lm(x2_2_di_22_6_bmp~group+age_at_study_participation, data=df01.2)
models <- list(m4, m4.1)
tab06 <- modelsummary(models,
             output = "huxtable",
             statistic = "p.value",
             stars = c("*" = .05, "**" = .01, "***" = .001),
             shape = term ~ model + statistic,
             sparse_header = TRUE,
             coef_rename = c("groupiPD" = "iPD",
                             "groupLRRK2 G2019S" = "LRRK2 G2019S",
                             "groupLRRK2 R1441G/C" = "LRRK2 R1441G/C",
                             "groupVPS35 D620N" = "VPS35 D620N",
                             "groupGBA" = "GBA",
                             "sexF" = "Sex",
                             "age_at_study_participation" = "Age"))

column_headers <- c("", "Model 1", "", "Model 2", "")
tab06 <-rbind(column_headers, tab06)
tab06[2,2] <- "Estimate"
tab06[2,3] <- "p-value"
tab06[2,4] <- "Estimate"
tab06[2,5] <- "p-value"

tab06 %>%
  huxtable::set_font_size(7)%>%
  huxtable::set_label("tab:tab06")%>%
  huxtable::set_caption("Summary of maximal model (1) and minimum sufficient model (2)
for 2,2'-di-22:6-BMP.")%>%
  huxtable::set_top_padding(2)%>%
  huxtable::set_bottom_padding(2)%>%
  huxtable::set_right_padding(2)%>%
  huxtable::set_left_padding(2)%>%
```

```r
huxtable::set_valign("middle")%>%
huxtable::set_header_rows(1:2, TRUE)%>%
huxtable::set_header_cols(1, TRUE)%>%
huxtable::style_headers(bold = TRUE)%>%
huxtable::merge_cells(1, 2:3)%>%
huxtable::merge_cells(1, 4:5)%>%
huxtable::set_align(everywhere, 2:5, "center")
```

Table 6: Summary of maximal model (1) and minimum sufficient model (2) for 2,2'-di-22:6-BMP.

| | **Model 1** | | **Model 2** | |
|---|---|---|---|---|
| | **Estimate** | **p-value** | **Estimate** | **p-value** |
| **(Intercept)** | 0.160 | 0.653 | 0.154 | 0.660 |
| **iPD** | 0.266 | 0.248 | 0.267 | 0.244 |
| **LRRK2 G2019S** | 1.539*** | 0.000 | 1.538*** | 0.000 |
| **LRRK2 R1441G/C** | 0.818** | 0.005 | 0.820** | 0.005 |
| **VPS35 D620N** | 1.347*** | 0.000 | 1.344*** | 0.000 |
| **GBA** | 0.044 | 0.879 | 0.048 | 0.866 |
| **Age** | 0.029*** | 0.000 | 0.029*** | 0.000 |
| **Sex** | -0.017 | 0.906 | | |
| **Num.Obs.** | 104 | | 104 | |
| **R2** | 0.541 | | 0.541 | |
| **R2 Adj.** | 0.507 | | 0.512 | |
| **AIC** | 240.5 | | 238.5 | |
| **BIC** | 264.3 | | 259.6 | |
| **Log.Lik.** | -111.231 | | -111.238 | |
| **RMSE** | 0.71 | | 0.71 | |
| *** p < 0.05, ** p < 0.01, *** p < 0.001** | | | | |

```r
par(mfrow = c(1,4))
plot(m4.1)
```

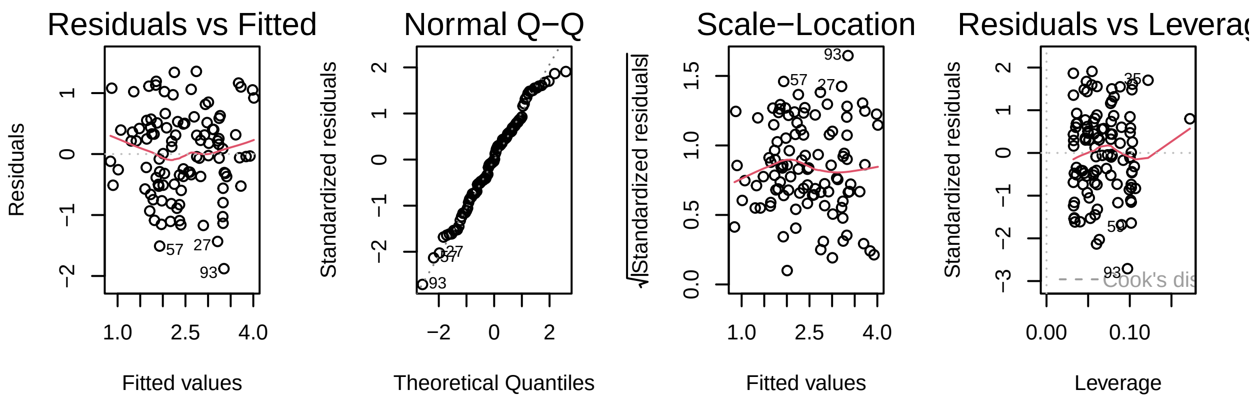

Figure 8: Residuals behaviour for the minimum sufficient model of 2,2'-di-22:6-BMP.

### 4 Session Information

```
sessioninfo::session_info()
```

```
## - Session info ---------------------------------------------------------------
##  setting  value
##  version  R version 4.2.1 (2022-06-23)
##  os       macOS Ventura 13.0.1
##  system   aarch64, darwin20
##  ui       X11
##  language (EN)
##  collate  en_US.UTF-8
##  ctype    en_US.UTF-8
##  tz       Europe/London
##  date     2022-11-22
##  pandoc   2.18 @ /Applications/RStudio.app/Contents/MacOS/quarto/bin/tools/ (via rmarkdown)
##
## - Packages -------------------------------------------------------------------
##  package       * version  date (UTC) lib source
##  abind           1.4-5    2016-07-21 [1] CRAN (R 4.2.0)
##  assertthat      0.2.1    2019-03-21 [1] CRAN (R 4.2.0)
##  backports       1.4.1    2021-12-13 [1] CRAN (R 4.2.0)
##  base64enc       0.1-3    2015-07-28 [1] CRAN (R 4.2.0)
##  bayestestR      0.12.1   2022-05-02 [1] CRAN (R 4.2.0)
##  bit             4.0.4    2020-08-04 [1] CRAN (R 4.2.0)
##  bit64           4.0.5    2020-08-30 [1] CRAN (R 4.2.0)
##  bookdown        0.28     2022-08-09 [1] CRAN (R 4.2.1)
##  broom           1.0.0    2022-07-01 [1] CRAN (R 4.2.0)
##  broom.helpers   1.8.0    2022-07-05 [1] CRAN (R 4.2.0)
##  car           * 3.1-0    2022-06-15 [1] CRAN (R 4.2.0)
##  carData       * 3.0-5    2022-01-06 [1] CRAN (R 4.2.0)
##  cellranger      1.1.0    2016-07-27 [1] CRAN (R 4.2.0)
##  checkmate       2.1.0    2022-04-21 [1] CRAN (R 4.2.0)
##  cli             3.3.0    2022-04-25 [1] CRAN (R 4.2.0)
##  codetools       0.2-18   2020-11-04 [1] CRAN (R 4.2.1)
##  colorspace      2.0-3    2022-02-21 [1] CRAN (R 4.2.0)
##  commonmark      1.8.0    2022-03-09 [1] CRAN (R 4.2.0)
##  cowplot         1.1.1    2020-12-30 [1] CRAN (R 4.2.0)
##  crayon          1.5.1    2022-03-26 [1] CRAN (R 4.2.0)
##  data.table      1.14.2   2021-09-27 [1] CRAN (R 4.2.0)
##  datawizard      0.5.1    2022-08-17 [1] CRAN (R 4.2.0)
##  DBI             1.1.3    2022-06-18 [1] CRAN (R 4.2.0)
##  dbplyr          2.2.1    2022-06-27 [1] CRAN (R 4.2.0)
##  digest          0.6.29   2021-12-01 [1] CRAN (R 4.2.0)
##  dplyr         * 1.0.9    2022-04-28 [1] CRAN (R 4.2.0)
##  DT            * 0.24     2022-08-09 [1] CRAN (R 4.2.0)
##  effectsize      0.7.0.5  2022-08-10 [1] CRAN (R 4.2.1)
##  ellipsis        0.3.2    2021-04-29 [1] CRAN (R 4.2.0)
##  emmeans         1.8.0    2022-08-05 [1] CRAN (R 4.2.0)
##  estimability    1.4.1    2022-08-05 [1] CRAN (R 4.2.0)
##  evaluate        0.16     2022-08-09 [1] CRAN (R 4.2.0)
##  fansi           1.0.3    2022-03-24 [1] CRAN (R 4.2.0)
##  farver          2.1.1    2022-07-06 [1] CRAN (R 4.2.0)
##  fastmap         1.1.0    2021-01-25 [1] CRAN (R 4.2.0)
##  flextable     * 0.7.3    2022-08-09 [1] CRAN (R 4.2.1)
##  forcats       * 0.5.2    2022-08-19 [1] CRAN (R 4.2.0)
```

```
##  fs              1.5.2    2021-12-08 [1] CRAN (R 4.2.0)
##  ftExtra       * 0.4.0    2022-04-20 [1] CRAN (R 4.2.0)
##  gargle          1.2.0    2021-07-02 [1] CRAN (R 4.2.0)
##  gdtools         0.2.4    2022-02-14 [1] CRAN (R 4.2.0)
##  generics        0.1.3    2022-07-05 [1] CRAN (R 4.2.0)
##  GGally        * 2.1.2    2021-06-21 [1] CRAN (R 4.2.0)
##  ggplot2       * 3.3.6    2022-05-03 [1] CRAN (R 4.2.0)
##  ggpubr        * 0.4.0    2020-06-27 [1] CRAN (R 4.2.0)
##  ggsci         * 2.9      2018-05-14 [1] CRAN (R 4.2.0)
##  ggsignif      * 0.6.3    2021-09-09 [1] CRAN (R 4.2.0)
##  glue            1.6.2    2022-02-24 [1] CRAN (R 4.2.0)
##  googledrive     2.0.0    2021-07-08 [1] CRAN (R 4.2.0)
##  googlesheets4   1.0.1    2022-08-13 [1] CRAN (R 4.2.0)
##  gt              0.7.0    2022-08-25 [1] CRAN (R 4.2.0)
##  gtable          0.3.0    2019-03-25 [1] CRAN (R 4.2.0)
##  gtsummary     * 1.6.1    2022-06-22 [1] CRAN (R 4.2.0)
##  haven           2.5.1    2022-08-22 [1] CRAN (R 4.2.0)
##  here          * 1.0.1    2020-12-13 [1] CRAN (R 4.2.0)
##  highr           0.9      2021-04-16 [1] CRAN (R 4.2.0)
##  hms             1.1.2    2022-08-19 [1] CRAN (R 4.2.0)
##  htmltools       0.5.3    2022-07-18 [1] CRAN (R 4.2.0)
##  htmlwidgets     1.5.4    2021-09-08 [1] CRAN (R 4.2.0)
##  httr            1.4.4    2022-08-17 [1] CRAN (R 4.2.0)
##  huxtable      * 5.5.0    2022-06-15 [1] CRAN (R 4.2.0)
##  insight         0.18.2   2022-08-10 [1] CRAN (R 4.2.1)
##  janitor       * 2.1.0    2021-01-05 [1] CRAN (R 4.2.0)
##  jsonlite        1.8.0    2022-02-22 [1] CRAN (R 4.2.0)
##  jtools        * 2.2.0    2022-04-25 [1] CRAN (R 4.2.0)
##  kableExtra    * 1.3.4    2021-02-20 [1] CRAN (R 4.2.0)
##  knitr           1.40     2022-08-24 [1] CRAN (R 4.2.0)
##  labeling        0.4.2    2020-10-20 [1] CRAN (R 4.2.0)
##  lattice         0.20-45  2021-09-22 [1] CRAN (R 4.2.1)
##  lazyeval        0.2.2    2019-03-15 [1] CRAN (R 4.2.0)
##  lifecycle       1.0.1    2021-09-24 [1] CRAN (R 4.2.0)
##  lubridate       1.8.0    2021-10-07 [1] CRAN (R 4.2.0)
##  magrittr        2.0.3    2022-03-30 [1] CRAN (R 4.2.0)
##  MASS            7.3-58.1 2022-08-03 [1] CRAN (R 4.2.0)
##  Matrix          1.4-1    2022-03-23 [1] CRAN (R 4.2.1)
##  modelr          0.1.9    2022-08-19 [1] CRAN (R 4.2.0)
##  modelsummary  * 1.0.2    2022-07-17 [1] CRAN (R 4.2.0)
##  multcomp        1.4-20   2022-08-07 [1] CRAN (R 4.2.0)
##  munsell         0.5.0    2018-06-12 [1] CRAN (R 4.2.0)
##  mvtnorm         1.1-3    2021-10-08 [1] CRAN (R 4.2.0)
##  officer         0.4.3    2022-06-12 [1] CRAN (R 4.2.0)
##  pander          0.6.5    2022-03-18 [1] CRAN (R 4.2.0)
##  parameters      0.18.2   2022-08-10 [1] CRAN (R 4.2.0)
##  performance     0.9.2    2022-08-10 [1] CRAN (R 4.2.1)
##  pillar          1.8.1    2022-08-19 [1] CRAN (R 4.2.0)
##  pkgconfig       2.0.3    2019-09-22 [1] CRAN (R 4.2.0)
##  plotly        * 4.10.0   2021-10-09 [1] CRAN (R 4.2.0)
##  plyr            1.8.7    2022-03-24 [1] CRAN (R 4.2.0)
##  purrr         * 0.3.4    2020-04-17 [1] CRAN (R 4.2.0)
##  R6              2.5.1    2021-08-19 [1] CRAN (R 4.2.0)
##  RColorBrewer    1.1-3    2022-04-03 [1] CRAN (R 4.2.0)
##  Rcpp            1.0.9    2022-07-08 [1] CRAN (R 4.2.0)
##  readr         * 2.1.2    2022-01-30 [1] CRAN (R 4.2.0)
```

```
##  readxl          1.4.1    2022-08-17 [1] CRAN (R 4.2.0)
##  reprex          2.0.2    2022-08-17 [1] CRAN (R 4.2.0)
##  reshape         0.8.9    2022-04-12 [1] CRAN (R 4.2.0)
##  rlang           1.0.4    2022-07-12 [1] CRAN (R 4.2.0)
##  rmarkdown       2.16     2022-08-24 [1] CRAN (R 4.2.0)
##  rprojroot       2.0.3    2022-04-02 [1] CRAN (R 4.2.0)
##  rstatix       * 0.7.0    2021-02-13 [1] CRAN (R 4.2.0)
##  rstudioapi      0.14     2022-08-22 [1] CRAN (R 4.2.0)
##  rvest           1.0.3    2022-08-19 [1] CRAN (R 4.2.0)
##  sandwich        3.0-2    2022-06-15 [1] CRAN (R 4.2.0)
##  scales          1.2.1    2022-08-20 [1] CRAN (R 4.2.0)
##  sessioninfo   * 1.2.2    2021-12-06 [1] CRAN (R 4.2.0)
##  snakecase       0.11.0   2019-05-25 [1] CRAN (R 4.2.0)
##  stringi         1.7.8    2022-07-11 [1] CRAN (R 4.2.0)
##  stringr       * 1.4.1    2022-08-20 [1] CRAN (R 4.2.0)
##  survival        3.4-0    2022-08-09 [1] CRAN (R 4.2.0)
##  svglite         2.1.0    2022-02-03 [1] CRAN (R 4.2.0)
##  systemfonts     1.0.4    2022-02-11 [1] CRAN (R 4.2.0)
##  tables          0.9.6    2020-09-22 [1] CRAN (R 4.2.0)
##  TH.data         1.1-1    2022-04-26 [1] CRAN (R 4.2.0)
##  tibble        * 3.1.8    2022-07-22 [1] CRAN (R 4.2.0)
##  tidyr         * 1.2.0    2022-02-01 [1] CRAN (R 4.2.0)
##  tidyselect      1.1.2    2022-02-21 [1] CRAN (R 4.2.0)
##  tidyverse     * 1.3.2    2022-07-18 [1] CRAN (R 4.2.0)
##  tzdb            0.3.0    2022-03-28 [1] CRAN (R 4.2.0)
##  utf8            1.2.2    2021-07-24 [1] CRAN (R 4.2.0)
##  uuid            1.1-0    2022-04-19 [1] CRAN (R 4.2.0)
##  vctrs           0.4.1    2022-04-13 [1] CRAN (R 4.2.0)
##  viridisLite     0.4.1    2022-08-22 [1] CRAN (R 4.2.0)
##  vroom           1.5.7    2021-11-30 [1] CRAN (R 4.2.0)
##  webshot         0.5.3    2022-04-14 [1] CRAN (R 4.2.0)
##  withr           2.5.0    2022-03-03 [1] CRAN (R 4.2.0)
##  xfun            0.32     2022-08-10 [1] CRAN (R 4.2.0)
##  xml2            1.3.3    2021-11-30 [1] CRAN (R 4.2.0)
##  xtable          1.8-4    2019-04-21 [1] CRAN (R 4.2.0)
##  yaml            2.3.5    2022-02-21 [1] CRAN (R 4.2.0)
##  zip             2.2.0    2021-05-31 [1] CRAN (R 4.2.0)
##  zoo             1.8-11   2022-09-17 [1] CRAN (R 4.2.0)
##
##  [1] /Library/Frameworks/R.framework/Versions/4.2-arm64/Resources/library
##
## ------------------------------------------------------------------------------
```
